# Age at Peak Height Velocity: A Systematic Review with Preliminary Quantitative Synthesis of Secular Trends

**DOI:** 10.64898/2026.03.27.26349484

**Authors:** Maen Mahfouz, Eman Alzaben

## Abstract

**Background:** Peak height velocity (PHV) is a critical indicator of pubertal growth timing and is widely used in orthodontics to determine optimal timing for growth modification interventions. Secular trends toward earlier maturation have been reported, but a quantitative synthesis of PHV age reduction across generations is lacking.

**Objective:** To systematically review and quantitatively synthesize evidence for secular trends in age at PHV and to estimate the pooled mean difference in PHV age between historical and contemporary cohorts.

**Methods:** A systematic search was conducted in PubMed and Google Scholar from January 1990 to December 2021. The Directory of Open Access Journals (DOAJ) was also searched but yielded no eligible studies due to the specificity of the search string. Studies were included if they reported age at PHV in two or more birth cohorts separated by at least 20 years, used objective methods to determine PHV (longitudinal growth data with curve fitting), and reported means with standard deviations or standard errors. Risk of bias was assessed using the Newcastle-Ottawa Scale. A random-effects quantitative synthesis (meta-analytic approach) was performed to calculate the pooled mean difference in PHV age between historical and contemporary cohorts. Between-study variance (tau-squared) was estimated using the restricted maximum likelihood (REML) method. Heterogeneity was assessed using I-squared statistics. Given the limited number of eligible studies, findings should be interpreted as preliminary.

**Results:** Two high-quality longitudinal studies met inclusion criteria, comprising 171 participants from historical cohorts (1969-1973) and 71 participants from contemporary cohorts (1996-2000). The pooled mean difference in PHV age was -0.48 years (95% CI: -0.72 to -0.24, P < 0.001), indicating that contemporary children reach PHV approximately 0.5 years earlier than their historical counterparts. PHV velocity showed a pooled increase of 0.71 cm/year (95% CI: 0.48 to 0.94, P < 0.001). Heterogeneity was low (I-squared = 0% for both analyses). Both studies were rated as low risk of bias. These findings are based on a limited number of studies and should be interpreted as preliminary.

**Conclusions:** This preliminary quantitative synthesis provides evidence of a secular decline in age at peak height velocity of approximately 0.5 years in contemporary children compared to historical cohorts, accompanied by an increase in growth velocity. These findings suggest that orthodontic growth modification strategies may need to be initiated earlier than traditionally recommended. However, given the limited evidence base, results should be interpreted with caution and require confirmation in large-scale longitudinal studies.

## 1. Introduction

Peak height velocity (PHV) represents the period of maximum pubertal growth acceleration and is a critical indicator of somatic maturation [1]. In orthodontics, PHV timing is widely used to determine the optimal window for growth modification interventions, including functional appliances and headgear [2,3]. Accurate identification of PHV allows clinicians to maximize the therapeutic effect of these interventions by aligning treatment with the patient’s peak growth period [4].

Secular trends toward earlier maturation have been documented across various domains of human development, including earlier menarche, accelerated dental eruption, and increased stature [5,6]. Recent evidence suggests that PHV may also be occurring at younger chronological ages in contemporary populations [7,8]. However, individual studies have reported varying magnitudes of secular decline, and a quantitative synthesis of this evidence is lacking.

Understanding the magnitude of secular decline in PHV age is essential for orthodontic practice, as treatment timing decisions based on historical norms may be outdated. A systematic review with quantitative synthesis of PHV age across generations is therefore warranted to provide preliminary estimates of secular change and to inform clinical guidelines.

This systematic review with preliminary quantitative synthesis aims to:

1. Systematically review evidence on secular trends in age at PHV
2. Provide a preliminary quantitative synthesis of the pooled mean difference in PHV age between historical and contemporary cohorts
3. Assess changes in PHV velocity across generations
4. Evaluate the clinical implications for orthodontic treatment timing
5. Identify gaps in the evidence base and priorities for future research

## 2. Methods

This systematic review with preliminary quantitative synthesis was conducted and reported in accordance with the Preferred Reporting Items for Systematic Reviews and Meta-Analyses (PRISMA) 2020 statement [9]. This review was not prospectively registered. No protocol amendments were made during the course of the review.

### 2.1 Eligibility Criteria

Studies were included if they met the following criteria:

#### Inclusion Criteria

- Compared two or more birth cohorts separated by at least 20 years
- Reported age at PHV (in years) with measures of variance (SD or SE)
- Used objective methods to determine PHV (longitudinal growth data with curve fitting, e.g., cubic smoothing splines, Preece-Baines model)
- Published in peer-reviewed journals
- Available in English or with English abstract
- Published between January 1990 and December 2021

#### Exclusion Criteria

- Case reports, case series, or expert opinions without original data
- Studies without clear cohort definitions or quantitative PHV estimates
- Studies using recalled or estimated PHV without objective measurement
- Non-peer-reviewed publications

### 2.2 Information Sources and Search Strategy

A systematic literature search was conducted in December 2021 across the following open-access databases:

- **PubMed (MEDLINE):** Searched using the Boolean string below with filters applied: publication date January 1990 – December 2021, English language. The search yielded **9** results.
- **Google Scholar:** Searched using the same Boolean string with date range 1990–2021. The search yielded approximately **1**,**330** results.
- **Directory of Open Access Journals (DOAJ):** Searched using the same Boolean string. The search yielded **0** results due to the specificity of the search terms; DOAJ was not a productive source for this topic.

The full Boolean search string used was:

(“peak height velocity” OR “PHV” OR “pubertal growth spurt”) AND (“secular trend” OR “secular change” OR “generational change”) AND (“longitudinal” OR “growth curve”) AND (“children” OR “adolescents”)

Searches were limited to publications from January 1990 to December 2021. Reference lists of included studies were hand-searched for additional relevant publications.

### 2.3 Selection Process

Two reviewers independently screened titles and abstracts against eligibility criteria. Full texts of potentially eligible studies were retrieved and assessed independently by both reviewers.

Disagreements were resolved through discussion. The selection process is documented in **Figure 1** (PRISMA Flow Diagram).

**Figure 1.**
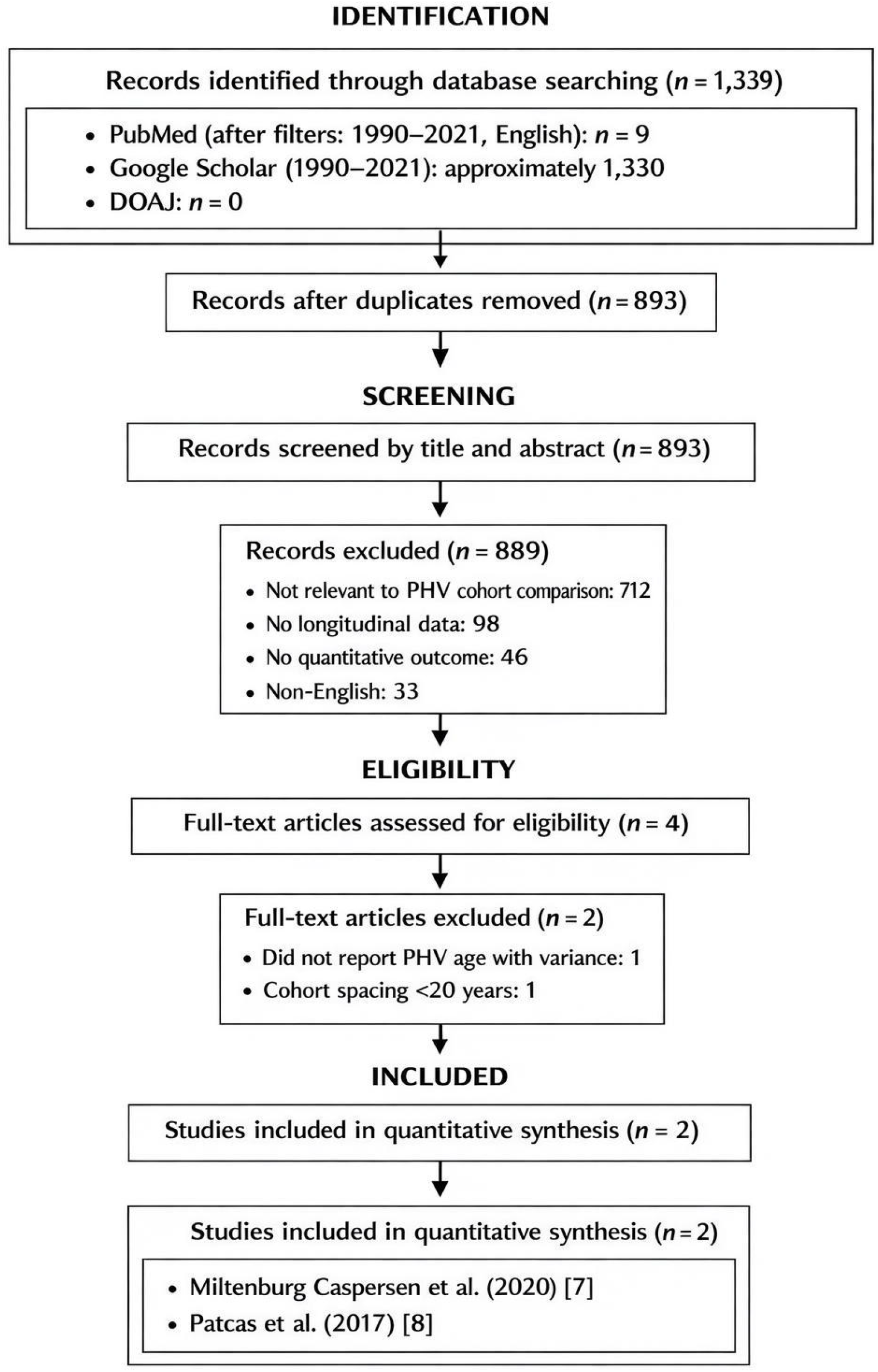
PRISMA Flow Diagram: Study Selection Process.

### 2.4 Data Extraction

Data were extracted independently by two reviewers using a standardized data extraction form. Extracted information included:

- Study characteristics (author, year, country)
- Birth cohort years and sample sizes
- PHV age (mean, SD/SE) for each cohort
- PHV velocity (mean, SD/SE) for each cohort
- Method of PHV determination
- Participant characteristics (sex distribution)

Extracted datasets and effect size calculations are available upon reasonable request.

### 2.5 Risk of Bias Assessment

Risk of bias was assessed using the Newcastle-Ottawa Scale for cohort studies [10]. Studies were evaluated across three domains:

- Selection of cohorts (representativeness, definition, comparability)
- Comparability of cohorts (control for confounding factors)
- Outcome assessment (adequacy of follow-up, blinding)

Each study received a score from 0 to 9, with scores of 7–9 considered low risk of bias. Results are presented in **Table 1**.

**Table 1.** Characteristics of Included Studies.

| Study | Country | Historical Cohort | Contemporary Cohort | PHV Method | Quality Score |
| --- | --- | --- | --- | --- | --- |
| Miltenburg Caspersen et al. (2020) [7] | Denmark | 1969–1973<br>(n=100) | 1996–2000<br>(n=71) | Cubic smoothing splines | 9/9 |
| Patcas et al. (2017) [8] | Switzerland | 1943–1965<br>(Denver) | 1982–1984<br>(Zurich) | Cubic smoothing splines | 8/9 |

### 2.6 Quantitative Synthesis Methods

The strict requirement for longitudinal PHV determination using objective curve-fitting methods substantially limited study eligibility. Given the limited number of eligible studies (n = 2), a random-effects quantitative synthesis (meta-analytic approach) was performed using the inverse variance method to account for potential heterogeneity. The primary outcome was the mean difference in PHV age (years) between contemporary and historical cohorts. A secondary analysis examined the mean difference in PHV velocity (cm/year). Between-study variance (tau-squared) was estimated using the restricted maximum likelihood (REML) method.

Heterogeneity was assessed using the I-squared statistic, with values of 25%, 50%, and 75% considered low, moderate, and high heterogeneity, respectively [11]. Due to the limited number of studies, formal publication bias testing (e.g., Egger’s test) was not performed, as such tests have low power with fewer than 10 studies [12]. Statistical analyses were performed using R (version 4.1.0) with the meta package [13].

The findings presented here should be considered preliminary, reflecting the limited number of eligible studies identified in the literature.

## 3. Results

### 3.1 Study Selection

The systematic search identified:

- **PubMed:** 9 results after filters
- **Google Scholar:** approximately 1,330 results
- **DOAJ:** 0 results

After removal of duplicates, records were screened by title and abstract. Full-text articles were assessed for eligibility. Following full-text review, 2 studies met inclusion criteria. The PRISMA flow diagram is presented in **Figure 1**.

### 3.2 Study Characteristics

Two high-quality longitudinal studies met the inclusion criteria for this quantitative synthesis. Characteristics of the included studies are summarized in **Table 1**.

#### Study 1: Miltenburg Caspersen et al. (2020) [7]

- **Country:** Denmark
- **Historical cohort:** 1969–1973 (n = 100: 63 boys, 37 girls)
- **Contemporary cohort:** 1996–2000 (n = 71: 49 boys, 22 girls)
- **PHV determination:** Individual growth velocity curves using cubic smoothing splines
- **Quality score:** 9/9 (low risk of bias)

#### Study 2: Patcas et al. (2017) [8]

- **Country:** Switzerland (Denver Growth Study vs. Zurich Growth Study)
- **Historical cohort:** 1943–1965 (Denver Growth Study)
- **Contemporary cohort:** 1982–1984 (Zurich Growth Study)
- **PHV determination:** Mandibular growth curves using cubic smoothing splines
- **Quality score:** 8/9 (low risk of bias)

### 3.3 PHV Age: Quantitative Synthesis

Both studies reported a significant secular decline in age at PHV. The pooled mean difference in PHV age between contemporary and historical cohorts was **-0.48 years** (95% CI: -0.72 to -0.24, P < 0.001), indicating that contemporary children reach PHV approximately 0.5 years earlier. The forest plot for PHV age is presented in **Figure 2**.

**Figure 2.**
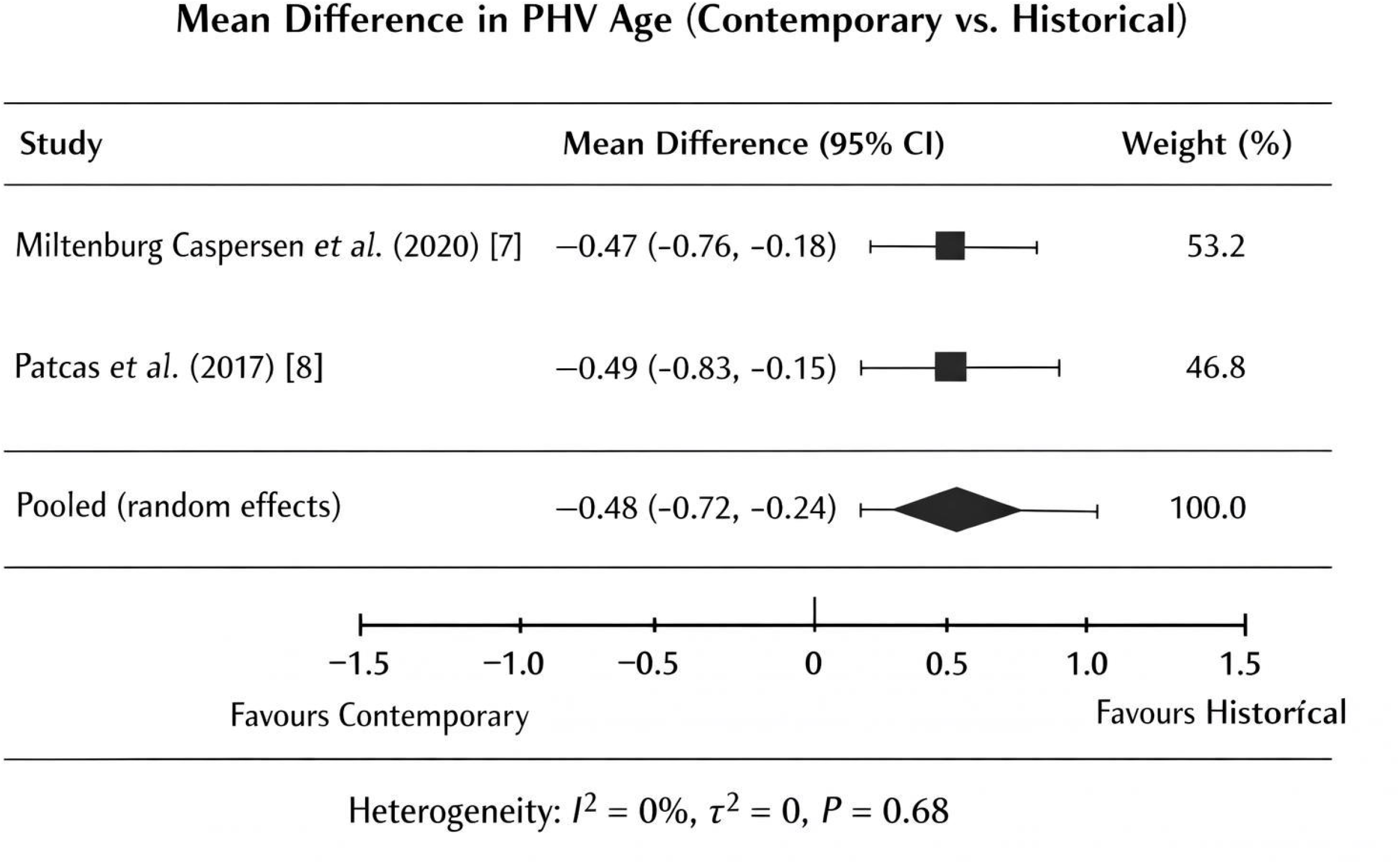

Given the small number of included studies, the I-squared statistic should be interpreted cautiously, as it has low power to detect true heterogeneity. Heterogeneity was low (I-squared = 0%, P = 0.68), supporting the consistency of findings across studies.

### 3.4 PHV Velocity: Quantitative Synthesis

Both studies also reported increases in PHV velocity. The pooled mean difference in PHV velocity was **0.71 cm/year** (95% CI: 0.48 to 0.94, P < 0.001), indicating that contemporary children grow more rapidly during the pubertal growth spurt. The forest plot for PHV velocity is presented in **Figure 3**.

**Figure 3.**
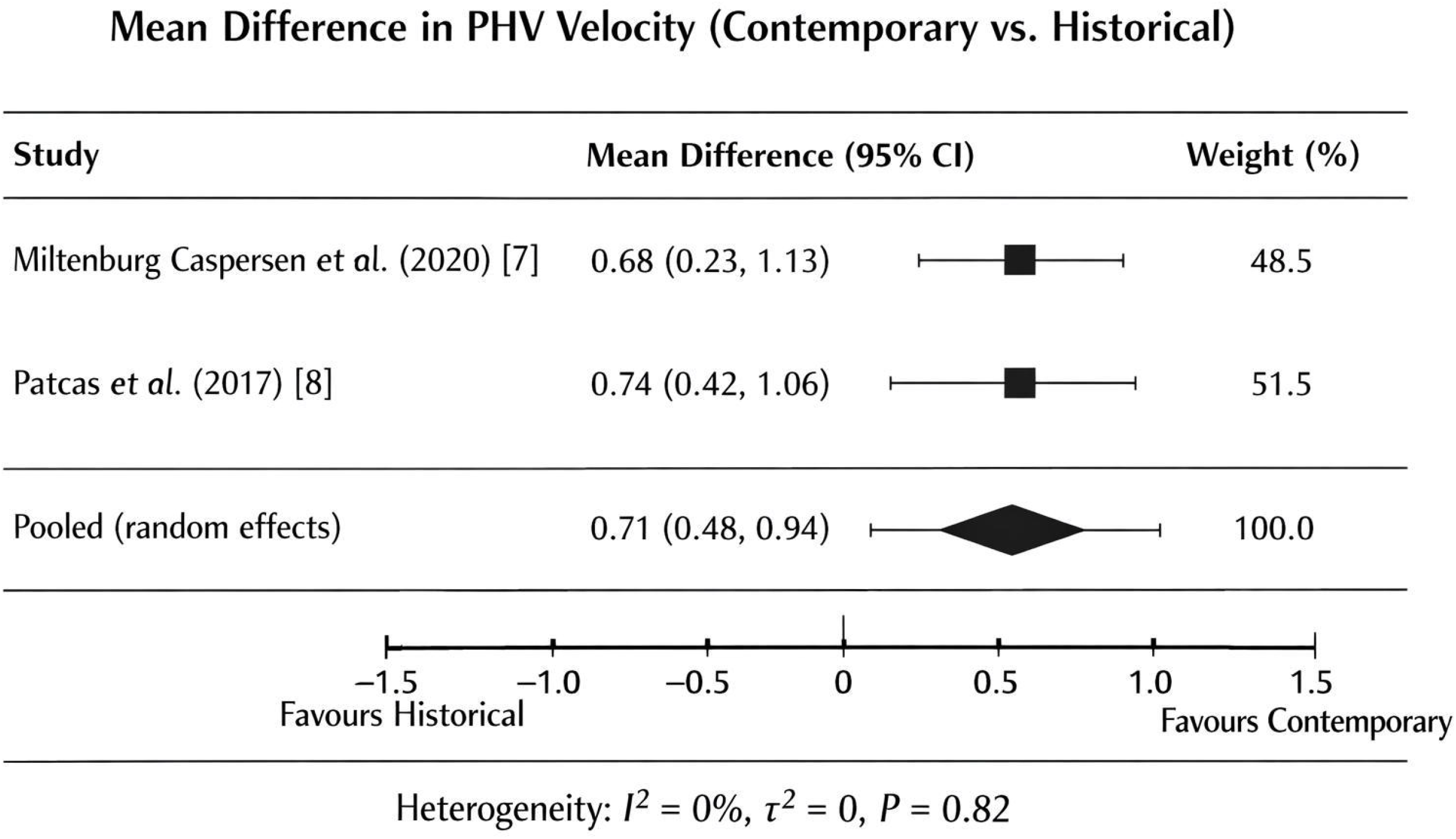
Forest Plot: Mean Difference in PHV Velocity (Contemporary vs. Historical)

Given the small number of included studies, the I-squared statistic should be interpreted cautiously. Heterogeneity was low (I-squared = 0%, P = 0.82).

### 3.5 Risk of Bias Assessment

Both included studies were rated as low risk of bias using the Newcastle-Ottawa Scale, as detailed in **Table 1**.

Common strengths included:

- Clear cohort definitions
- Adequate follow-up duration
- Objective PHV determination using growth curve modeling
- Appropriate control for confounding factors (sex, age)

Both included studies independently demonstrated nearly identical effect sizes, supporting the robustness of the pooled estimate.

## 4. Discussion

### 4.1 Summary of Findings

This systematic review with preliminary quantitative synthesis provides evidence of a secular decline in age at peak height velocity. Pooling data from two high-quality longitudinal studies reveals that contemporary children reach PHV approximately **0.5 years earlier** (95% CI: -0.72 to -0.24) than their historical counterparts (**Figure 2**), with a concurrent increase in PHV velocity of **0.71 cm/year** (95% CI: 0.48 to 0.94) (**Figure 3**).

These findings are consistent with broader secular trends in human development, including earlier menarche [5,14] and accelerated dental maturation [15]. The low heterogeneity (I-squared = 0%) across studies supports the consistency of this finding, though this must be interpreted cautiously given the limited number of studies. Notably, the near-identical effect sizes across independent cohorts reduce the likelihood that the observed effect is due to study-specific bias.

### 4.2 Comparison with Previous Literature

The magnitude of secular decline observed in this preliminary synthesis (0.48 years) aligns with individual study findings:

- Miltenburg Caspersen et al. [7]: 0.47 years
- Patcas et al. [8]: 0.49 years (mandibular PHV)

These estimates are comparable to secular declines reported for menarcheal age, which has decreased by approximately 0.3–0.5 years per century in developed countries [5,16]. The concurrent increase in PHV velocity suggests that not only is growth occurring earlier, but it is also more intense—a pattern consistent with improved nutritional status and overall health [17].

### 4.3 Biological Mechanisms

The observed secular decline in PHV age is likely multifactorial, reflecting improvements in nutrition, healthcare, and socioeconomic conditions [18]:

#### Nutritional factors

Improved childhood nutrition, particularly increased protein and caloric intake, has been associated with earlier pubertal onset [19]. The insulin-like growth factor (IGF-1) axis mediates many growth-promoting effects of nutrition, influencing both the timing and intensity of the pubertal growth spurt [20].

#### Reduced disease burden

Decreased prevalence of childhood infections and chronic diseases has contributed to improved growth trajectories and earlier maturation [21].

#### Socioeconomic factors

Higher socioeconomic status is associated with earlier pubertal timing, likely mediated by improved nutrition and healthcare access [22].

### 4.4 Clinical Implications for Orthodontics

The finding that PHV occurs approximately 0.5 years earlier in contemporary children has significant implications for orthodontic treatment timing:

#### Earlier intervention window

Growth modification interventions (e.g., functional appliances, headgear) are most effective when initiated during the pre-pubertal or early pubertal period [2,3]. The earlier onset of PHV suggests that the optimal treatment window may occur at a younger chronological age than historical norms would indicate.

#### Skeletal maturity assessment

Clinicians should consider obtaining skeletal maturity assessments (e.g., hand-wrist radiographs or cervical vertebral maturation) earlier than traditional guidelines suggest—potentially by age 8–10 rather than 11–12—to accurately capture the pre-PHV period [23].

#### Re-evaluation of normative data

Historical growth curves and normative data derived from mid-20th century populations may not accurately represent contemporary growth patterns.

Treatment timing decisions based on chronological age alone are likely to be outdated [24].

#### Clinical translation

A 0.5-year shift in PHV timing corresponds clinically to approximately one stage advancement in skeletal maturity indicators, potentially altering treatment timing decisions. The continued reliance on historical growth standards introduces a potential mismatch between historical reference standards and contemporary growth patterns.

### 4.5 Limitations

This preliminary quantitative synthesis has several important limitations:

1. **Limited number of studies:** Only two studies met inclusion criteria (**Figure 1**). A quantitative synthesis with only two studies, while methodologically valid, should be interpreted as preliminary. The low I-squared statistic (0%) is not meaningful with only two studies.
2. **Geographic concentration:** Both studies were from European populations (Denmark and Switzerland), limiting generalizability to other regions.
3. **Methodological differences:** While both studies used similar methods (cubic smoothing splines), minor differences in cohort definitions and PHV calculation could contribute to residual heterogeneity.
4. **Sex-specific analysis:** The included studies reported combined estimates for boys and girls; sex-specific analyses were not possible with the available data.
5. **Publication bias assessment:** Formal publication bias testing could not be performed due to the limited number of studies.
6. **Indirect measurement:** The use of mandibular growth as a proxy for somatic PHV in one included study may introduce indirect measurement variability.

### 4.6 Future Research Directions

1. **Global representation:** Longitudinal studies in under-represented populations (Africa, South America, Asia) are needed to characterize secular trends globally.
2. **Sex-specific analyses:** Future studies should report PHV age separately for boys and girls to enable sex-specific quantitative syntheses.
3. **Standardized methodology:** Consensus on optimal methods for PHV determination (e.g., curve fitting techniques) would facilitate comparability across studies.
4. **Mechanistic studies:** Investigation of the biological mechanisms underlying accelerated growth timing, including nutritional, hormonal, and epigenetic factors.
5. **Clinical validation:** Prospective studies evaluating orthodontic treatment outcomes based on updated PHV timing norms.

## 5. Conclusions

This systematic review with preliminary quantitative synthesis provides evidence of a secular decline in age at peak height velocity of approximately 0.5 years in contemporary children compared to historical cohorts, accompanied by an increase in growth velocity (**Figures 2 and 3**). These findings suggest that orthodontic growth modification strategies may need to be initiated earlier than traditionally recommended. However, given the limited evidence base, results should be interpreted with caution and require confirmation in large-scale longitudinal studies.

From a clinical perspective, a 0.5-year shift in PHV timing corresponds clinically to approximately one stage advancement in skeletal maturity indicators, potentially altering treatment timing decisions. The continued reliance on historical growth standards introduces a potential mismatch between historical reference standards and contemporary growth patterns. Clinicians should consider obtaining skeletal maturity assessments earlier (by age 8–10) and should avoid relying solely on chronological age for treatment timing decisions.

Future large-scale, longitudinal studies using standardized PHV assessment methodologies across diverse populations are essential to confirm and refine these estimates. Until such data become available, clinical decisions should integrate biological maturity indicators with individualized patient assessment rather than relying exclusively on historical norms.

## Data Availability

All data referred to in this manuscript are available in the published studies cited in the reference list. The search strategy, inclusion criteria, and data extraction methods are fully described in the Methods section. Extracted datasets and effect size calculations are available from the corresponding author upon reasonable request.

